# Mapping infectious disease mortality onto the within-host dynamics of pathogen growth

**DOI:** 10.1101/2020.06.20.20134205

**Authors:** Tim O’Sullivan, Kristofer Wollein Waldetoft, Sam P. Brown

## Abstract

Infectious diseases remain a major cause of global mortality, yet basic questions concerning the relationship between within-host processes governing pathogen burden (pathogen replication, immune responses) and epidemiological patterns of mortality remain obscure. In standard epidemiological models of bacterial and viral infections (‘compartmental models’), pathogen-induced mortality is described as a constant risk factor, unchanging with time since infection or the dynamics of the pathogen population inside the host. Here we review experimental data illustrating the failure of this assumption, with multiple infection models highlighting an accelerating risk of death with time since infection. Next, we show that the dynamics of infection-mediated death are phenomenologically similar to the dynamics of all-causes death, captured by an exponentially increasing instantaneous mortality or ‘hazard’ (a Gompertz function). We then sketch a series of within-host pathogen models and highlight that multiple models are consistent with existing Gompertz-like infection mortality data, including: (1) exponential pathogen growth and a linear mapping between pathogen density and host hazard, and (2) linear pathogen growth and an exponential mapping between pathogen density and host hazard. We review critical experimental questions that our work highlights, requiring additional non-invasive data on pathogen burden throughout the course of infection, and end with a discussion on the unpicking of the mechanistic and dynamical basis of accelerating mortality risk during the course of human infections.

## Introduction

Infectious disease research operates across multiple scales, from cell biologists studying the molecular interactions between pathogen and host cells, through to epidemiologists studying the trajectories of infection case counts through populations of hosts. The breadth of infection research is a testament to the enduring importance of infections to humanity and provides an incredible resource to infection researchers working across these scales. Yet the specialization of research has led to a relative disconnect between the within-host focus of biomedical research and the among-host focus of epidemiological research. In particular, we focus in this study on a disconnect over the analysis of a key infection outcome – life or death.

On an epidemiological scale, the standard model framework for bacterial and viral (‘microparasite’) infections is known as ‘compartmental’ models, which structure host populations into compartments of ‘susceptible’ (S) and ‘infected’ (I) classes [1]. Under these models, individuals in the S class can transition into the I class (following contact with an I individual), and then exit the I class following pathogen clearance or death (Figure 1A). A key simplification in these models is that the mortality rate of an individual jumps from a baseline rate µ (while uninfected) to µ + *α* while infected (*α* capturing the additional pathogen-induced mortality), and returning immediately to µ when and if the infection is cleared (Figure 1B). A consequence of this modeling approach is that probability of survival will follow an exponential decay through time since infection (Figure 1C), reflecting the constant and elevated risk posed by infection. This constant mortality assumption has been flagged before as problematic [2–4] and is commonly relaxed in epidemiological models with very heterogeneous infection burdens, such as macroparasitic worm infections [5,6]. Here we focus on microparasite infections and ask first what are the empirical patterns of mortality following experimentally induced infections, and then develop statistical and mathematical models of dynamical mortality in light of these data.

**Figure 1.**
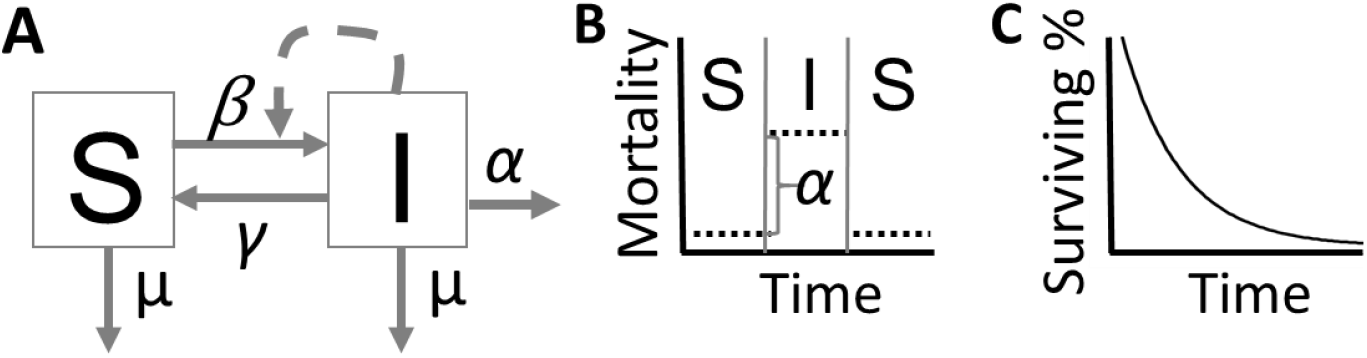
A schematic summary of mortality assumptions in a compartmental SIS epidemiological model. (A) An SIS compartmental model. Individuals are allocated to either the S or I class, dependent on whether susceptible or infected. Transitions among states are governed by coefficients capturing transmission (β), recovery (γ) and mortality (α). (B) The transitions between S and I states in model (a) imply instantaneous switches in mortality, between a background rate µ and an enhanced rate µ + α. (C) The constant (and elevated) rate of instantaneous mortality during infection implies that among the infected class, survivors (in the absence of subsequent infections or recoveries) will decay exponentially with time since infection.

## Results

To assess the validity of standard epidemiological assumptions (Figure 1), we gathered data that represent a phylogenetically diverse range of microparasites and metazoan hosts [7–10], including a fruit fly / *Listeria monocytogenes*, mouse / *Trypanosoma brucei*, waxmoth / *Pseduomonas aeruginosa*, and nematode / *Staphylococcus aureus* models. These infection models represent many different mechanisms of host immunity and pathogen virulence, and thus capture a large diversity of parasite-host interactions. Each dot in Figure 2 illustrates the proportion of surviving individuals at time *t* since experimental infection. In these experiments, age-controlled cohorts were infected at defined times and with a defined pathogen dose or exposure. In the fruit fly and waxmoth models, hosts were infected with a known number of colony forming units (CFU) of the bacterial pathogen. For the mouse model, hosts were infected with a known number of parasites, cultured from immunosuppressed mouse hosts. Nematodes were infected with a typical slow killing assay [11], which involved a discrete exposure time of feeding on the bacterial pathogen (i.e. in *C. elegans* and allied bacterivorus species, infections are the results of post-feeding colonization, not injection).

**Figure 2.**
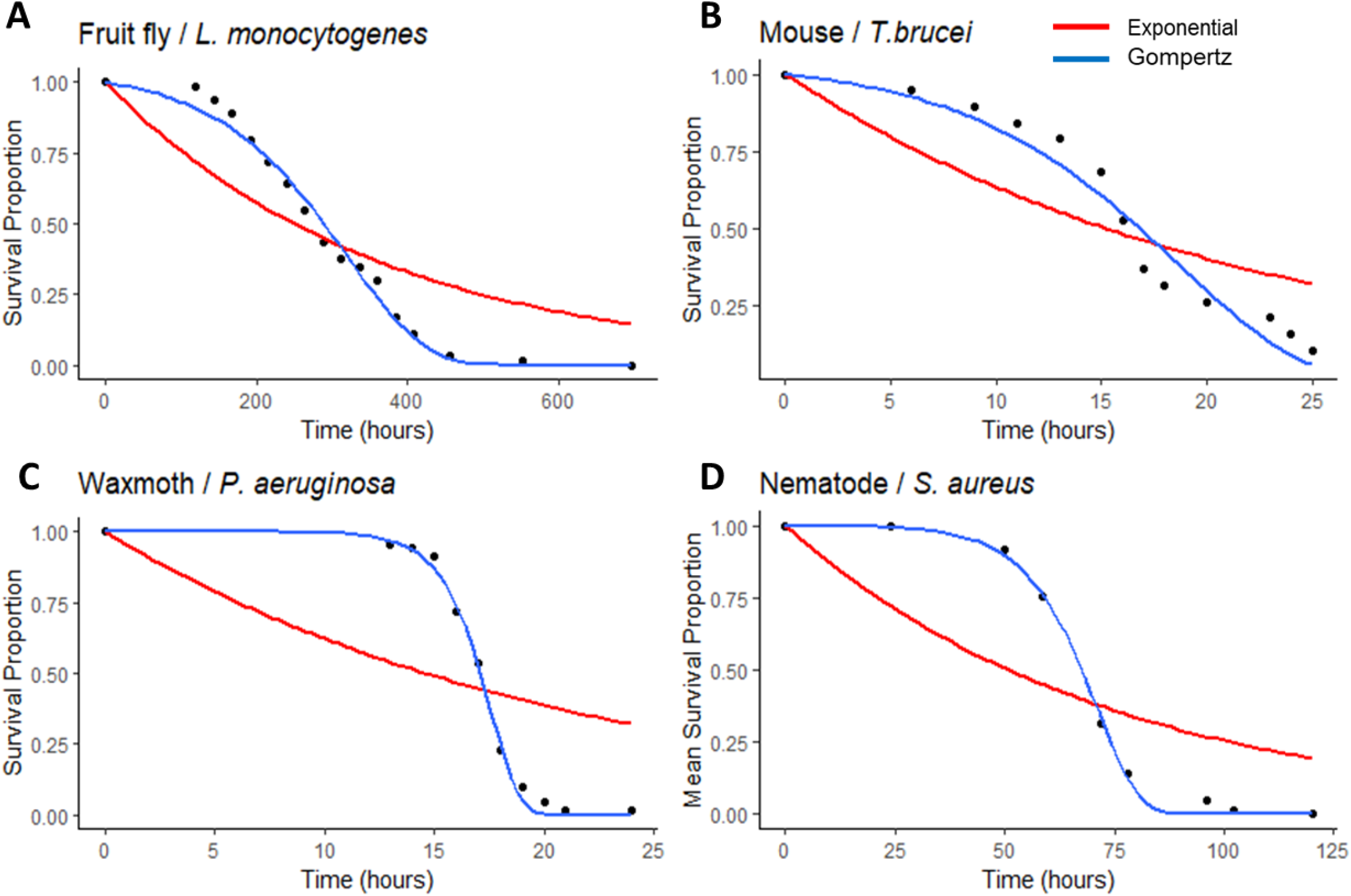
Survival data for four host-microparasite model systems indicate the failure of the constant mortality assumption. Black dots represent experimental data from (A) a fruit fly/ Listeria model (N=64, [7]), (B) a mouse/trypanosome model (N=19, [8]), (C) a waxmoth/*P. aeruginosa* model (N=71, [9]), and (D) a nematode/*S. aureus* model (N=120, [10]). The red lines represent exponential model fits (constant mortality rate), the blue lines represent a Gompertz model fit, which allows for accelerating mortality in time. Model fits were made using a nonlinear least squares method in R [22]. Parameter estimates and model comparisons are in Table 1.

Using the entire survival data, we next fitted an exponential model for each system (red lines, Figure 2). This corresponds to a constant mortality rate for each host-parasite combination, as is assumed in the epidemiological compartmental modeling approach discussed above (Figure 1). On even a cursory examination, the data are not consistent with an exponential decline in survivorship. All the survivorship curves systematically deviate from the best fit exponential with higher survivorship early in the infection, and lower survivorship late in the infection. In other words, the data support an increasing risk of death as the infection proceeds.

**Table 1.** Comparison of exponential and Gompertz model fits.

| <u>Data</u> | <u>Model</u> | <u>AIC</u> | <u>Fitted Parameters</u> |
| --- | --- | --- | --- |
| Fruit fly / *L.monocyto*genes [7] | Exponential | -7.21532 | $\lambda = 0.002778485$ |
| | Gompertz | -56.16302 | $b = 0.0004192727$ , $a = 0.0098624234$ |
| Mouse / *T. brucei* [8] | Exponential | -5.217522 | $\lambda = 0.045424$ |
| | Gompertz | -27.34543 | $b = 0.007434867$ , $a = 0.166813158$ |
| Waxmoth / *P. aeruginosa* [9] | Exponential | 9.718088 | $\lambda = 0.047421$ |
| | Gompertz | -42.53678 | $b = 1.150947e-06$ , $a = 0.7620354$ |
| Nematode / *S. aureus* [10] | Exponential | 4.020253 | $\lambda = 0.013633$ |
| | Gompertz | -39.09753 | $b = 6.250612e-05$ , $a = 0.1044699$ |

### Pathogen-induced death as a process of accelerated aging

This pattern of increasing instantaneous mortality or ‘hazard’ through time is familiar from the aging literature. In a human context, the instantaneous risk of death increases approximately exponentially, doubling approximately every 7 years [12]. In controlled animal aging experiments, the same statistical patterns are observed, with the absolute rates varying with the animal [13–15], their genetics [16], and the environment [17,18]. Inspired by this connection, we decided to test one of the most common models from the aging literature, the Gompertz [19]. Briefly, the two parameter Gompertz mortality function is defined by:

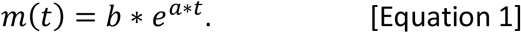

When *a* = 0, the Gompertz recovers the exponential distribution with a constant mortality rate *b*. Increasing *a* leads to an increasing acceleration of instantaneous mortality with time [20,21]. We used a nonlinear least squares method [22] in the statistical software R [23] to fit these distributions to the experimental data, and observe that models of accelerated mortality (akin to aging) fit the survival data better than the exponential model (Figure 2), but at the expense of an additional parameter. To assess model fit while accounting for parameter number, we used an information criterion approach, as presented in Table 1. The results clearly show the extra parameter in the Gompertz model is well justified, supporting the conclusion that in these diverse models, the risk of death is accelerating following the initiation of infection – analogous to the acceleration in mortality across entire lifespans, known as aging.

However, while experimental infection and aging give rise to similar patterns of survival, there is a difference in underlying process. In the case of the experimental infection models illustrated above, we have a clear and singular cause of death – the pathogen (uninfected controls showed minimal mortality across the timescale of observation, specifically, survivorship in control (uninfected) cohorts at the final time point ranged from ∼80% (waxmoth) to 100% (nematodes) [7–10]. This causal clarity offers a window into the study of mortality, as we have a potential internal currency (pathogen dynamics) to map onto the changing risk of death.

### Pathogen-driven models of accelerating mortality

The observation of an approximately exponential increase in risk (*i*.*e*., a Gompertz function) suggests one simple mechanistic model: the instantaneous mortality is linearly proportional to the instantaneous burden of an exponentially expanding pathogen population. We can break this down into two sets of assumptions. First, we assume that within-host pathogen density *p*(*t*) is growing exponentially, *p*(*t*) = *p*_0_ *e*^*rt*^, with dynamics governed by initial inoculum *p*_0_ and an exponential pathogen growth rate *r*. Given an acute infection, this is a plausible assumption and commonly made in ‘nested’ epidemiological models [2]. Next, we assume that instantaneous mortality *m*(*t*) is linearly proportional to bacterial burden, with mortality coefficient *v*, that is *m*(*t*) = *v* ∗ *p*(*t*). This again is a plausible assumption, capturing the notion that higher pathogen burdens are more dangerous. Putting these pieces together, we arrive at an instantaneous mortality function that is driven by pathogen demography, and is identical to the Gompertz function (given *b* = *v* ∗ *p*0 and *a* = *r*);

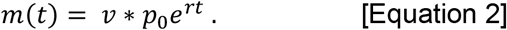

In Figure 3, we focus on the mouse / *T. brucei* infection model (Kaboré, Figure 2B), and use our fitted Gompertz function (Table 1) plus independent estimates of inoculum density (*p*_0_ = 5 * 10^4^) to parameterize our mechanistically explicit model in Equation 2. From this exercise, we find *r* = *a* ∼ 0.167/*hr* and per-capita virulence *v* = *b*/*p*0 ∼ 1.5 ∗ 10 − 7 / (*cells* ∗ *hr*). Plotting the implied pathogen growth through time (blue line, Figure 3) the model therefore predicts that by 25 hours, when the majority of hosts are dead, pathogen density has reached approximately 3 ∗ 10^7^, a plausible density in murine-trypanosome infection models [24,25]. While the model assumptions of exponential growth and a linear mapping appear plausible, and the resulting Gompertz equation fits the data above, we must heed the caution that multiple mechanistic processes can be consistent with a single statistical pattern [26,27]. In the following section we explore a number of scenarios that are consistent with plausible within-host processes and also generate the observed acceleration in risk during the course of infection.

**Figure 3.**
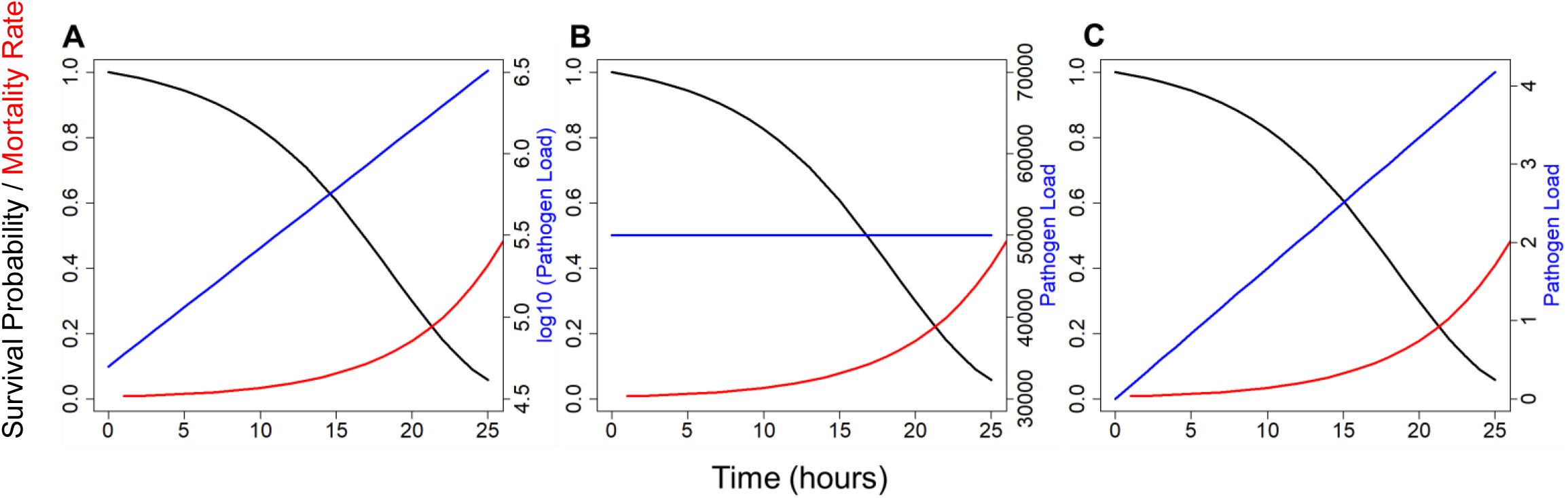
Three distinct within-host models that are consistent with the observed survival data for the mouse / *T. brucei* system. A) Exponential pathogen growth with a linear mapping from pathogen density to instantaneous mortality. B) constant pathogen load post infection, with independent and exponentially increasing mortality. C) linear pathogen growth with exponential mapping to mortality. Model fits are to the data in Figure 2a.

Data on the within-host dynamics of infection are sadly limited, but in some studies there is little change through time in pathogen density [28], and in some cases there are even appreciable declines – albeit from deliberately high inocula [29]. In light of these data, we could conservatively assume that within-host dynamics are relatively constant, *i*.*e*., *p*(*t*) = *p*_0_. From this standpoint, we can still return to a Gompertz mortality function by assuming *m*(*t*) = *b* ∗ *e*^*a* ∗ *t*^, *i*.*e*., the acceleration in risk is independent of the demography of the pathogen. Figure 3B illustrates this model, again fitted to the Kaboré et al. data [8]. In this case, we can use observed *p*0 data to constrain pathogen load, but by assumption this has no mechanistic connection to changing mortality.

In a third avenue of approach, we assume a linear expansion of the pathogen following infection at rate *g*, (*p*(*t*) = *p*_0_ + *gt*). In this case, we can again return to a Gompertz function in the limit of *p*0 −> 0, given that mortality is an exponential function of pathogen density *m*(*t*) = *v* ∗ *e*^*p*(*t*)^. By substation, we can now connect within-host pathogen growth to population mortality by *m*(*t*) = *v* ∗ *e*^*g*(*t*)^. Applying this function to the Kong data (Figure 3C) we see that while we fit the survival data, the implied within-host behavior is not plausible in this case, implying pathogen loads that remain well below the observed *p*_0_. Finally, in Figure S1 we take the same 3 models illustrated in Figure 3 for the mouse/trypanosome system and apply to the other 3 experimental model systems. While we see different implied within-host behaviors with differing plausibilities, the main take home from this analysis is that inferring within-host behavior purely from mortality trajectories is an under-determined problem. In short, we need to look inside hosts to directly observe pathogen dynamics.

## Discussion

In contrast to the assumptions in standard epidemiology models (Figure 1), our analysis shows that the instantaneous rate of mortality increases exponentially during the course of infection, in 4 diverse experimental models of infection (Figure 2). This exponential increase in risk is a hallmark of aging in humans [30] and other organisms [31], and is phenomenologically described by the two parameter Gompertz equation (Table 1).

Unlike the aging literature, however, we have in the case of acute lethal infections a clear causal currency – the bacterial (Figure 1; A, C, and D) or protozoan (B) pathogen. Building on this causal connection, we next outline potential mechanistic paths between the within-host dynamics of the pathogen and the observed acceleration in mortality (Figure 3). Our analysis demonstrates that multiple causal processes are consistent with the observed survival data, illustrating that the observation of accelerating mortality alone is insufficient to infer the underlying process of mortality.

Our mechanistic analyses highlight two sets of assumptions that together combine to define instantaneous mortality, *m*(*t*), in terms of pathogen dynamics *p*(*t*). First, we need to define the nature of pathogen growth *p*(*t*). In Figure 3 we consider exponential (Figure 3A), linear (Figure 3C) or no growth (Figure 3B). Other reasonable choices include logistic growth, or even declines from a high inoculum, as observed in some experimental models with large challenge inoculations [28]. Second, we need to define how pathogen dynamics *p*(*t*) shape instantaneous mortality *m*(*t*). In Figure 3 we consider a linear mapping (Figure 3A), exponential mapping (Figure 3C) or *m*(*t*) independent of *p* (Figure 3B). Again, other choices could be made, for instance a mapping incorporating infection history, *m*(*t*) ∼ *∫ p*(*t*) *dt*. For each of these sets of assumptions on the form of *p*(*t*) and *m*(*t*), we next review possible data sources to constrain and inform our model assumptions.

To address the challenge of measuring within-host pathogen dynamics *p*(*t*), broadly two paths are available. The first and simplest path is destructive sampling, where a large cohort is tracked during the course of infection, and sample individuals are taken at intervals and sacrificed to estimate pathogen burden at time *t*, e.g. by grinding up the host or host tissue and plating on selective media for a defined bacterial pathogen [32,33]. Destructive sampling is simple in outline, but raises challenges of individual host heterogeneity (data is gathered across hosts) and the requirement for very large cohort sizes in order to gather data late in the progression of infection [34,35]. A second path is to track infection burden non-destructively via repeat measures from the same population of hosts. While requiring fewer hosts to produce workable data, the requirement to minimize invasive observational effects can become a limitation [36]. One attractive route is to use fluorescent or luminescent pathogen strains, that can be imaged within a host non-invasively [37]. This path has been taken in diverse animal models, from mice [38] to nematodes [39], although calibration of infection burden is a concern – typically these reporter construct studies are more focused on location of infection [28,40] rather than estimating burden. In the absence of light-based reporters, other routes are possible via the sampling of peripheral fluids, e.g. for malaria [41,42]. However the risk of observer effects are higher in these cases, as attempting to sample from moribund hosts one may inadvertently accelerate mortality, thereby warping the relationship between pathogen burden and death [43,44].

Turning to the mapping function, *m*(*t*) = *f*(*p*(*t*)), some evidence can be gleaned from experiments varying inocula [45–47], but without tracking subsequent dynamics of the pathogen within the host, this approach is limited. To empirically estimate mapping functions will require empirical measures of instantaneous mortality *m*(*t*) (from survival data, e.g., Figure 3), together with measures of pathogen density, *p*(*t*), as outlined in the previous paragraph. In future work, we aim to pursue this agenda using an experimental *C. elegans* infection system.

In our current work, we focused on diverse infection models, with divergent modes of pathogen replication and host immune control. Despite this biological diversity, we witnessed a similar phenomenological pattern of accelerating risk. We note however that while our results are illustrative of diverse infection systems, they do not represent a formal meta-analysis of infection data, and we aim to conduct a systematic literature review in future work.

Returning to the epidemiological focus of our introduction, what are the epidemiological implications of a failure of the constant mortality assumption in compartmental models (Figure 1)? In contrast to our study of synchronized cohorts of experimentally induced infections (Figure 2), in most natural epidemiological contexts, the onset of infections are un-synchonised, and the age of infection (*t* in our analyses above) is unknown. In this context, the standard SIS model framework is often reasonable, and when the model produces dynamics that are ‘too fast’ due to the sudden jump from S to I, simple fixes are available such as through the addition of a latent or ‘exposed’ class in SEIR (Susceptible-Exposed-Infected-Recovered) models to delay epidemiological progress [48]. Our approach raises the potential for incorporating age of infection (*t*) and pathogen dynamics *p*(*t*) into analyses, a topic which has been pursued under the banner of ‘nested’ epidemiological models [49,50]. While previous work on nested epidemiology has relied on ‘plausible’ models of *p*(*t*), e.g., Antia et al. 1994, Gilchrist & Sasaki 2002 [51,52], we caution that multiple within-host processes can be consistent with epidemiological data (Figure 3). What we now require are studies to empirically determine the within-host dynamics of infection and their mapping to mortality. With this data we will be able to further constrain models and explore new avenues for intervention strategies that are conditioned on the progressive state of infectious disease.

## Data Availability

The data used for this study were drawn from a mix of studies that were either publicly available or personally requested from their respective authors. Data, as well as R scripts will be made available upon request.

